# Combining Randomized and Observational Data: The Case of Postnatal Prophylaxis Against HIV

**DOI:** 10.64898/2026.10.15.26365628

**Authors:** Dwight E. Yin, Paul N. Zivich, Mark J. Giganti, Danni Wu, Jessie K. Edwards, Stephen R. Cole

## Abstract

The efficacy of current antiretroviral interventions for prevention of vertical HIV transmission is both a triumph and a challenge for future efficacy studies of novel agents, including long-acting antiretroviral agents. Given the effectiveness of current antiretroviral interventions, standard randomized controlled trials to assess novel interventions targeted towards further reducing vertical HIV transmission may be infeasible due to required sample sizes. To address this study design challenge, we propose a novel augmented clinical trial design, where a randomized trial with an active comparator is augmented with a concurrent observational cohort to supplement the standard-of-care arm. The proposed design and analysis can overcome sampling biases, and an associated diagnostic can evaluate potential biases from unmeasured covariates. The proposed trial design is illustrated with an example and studied further using a simulation study. In the illustrative example, an augmented trial with a 3:1 allocation ratio in the trial component yielded an unbiased treatment effect estimate, as compared to the trial component alone, and the standard error was reduced substantially. Although the total sample size of the augmented trial was larger than a similarly powered 1:1 standard trial, the sample size of the augmented trial’s trial component was half the size of the standard trial and within the range of historically similar trials, thereby providing both feasibility and potential cost advantages. Although limitations with this design remain, many of these limitations can be mitigated by careful study planning and coordination. Augmented trials provide a potentially applicable solution to studying the efficacy of long-acting antiretrovirals for infant postnatal prophylaxis against HIV.

## Introduction

The standard-of-care to prevent vertical (i.e., mother-to-child) HIV transmission, maternal HIV treatment with antiretroviral therapy coupled with daily oral infant antiretroviral prophylaxis, can reduce transmission risk to approximately 1 percent.^1–8^ Nonetheless, an estimated 120,000 infants acquire HIV each year.^9^ A preponderance of these new cases occur via breastfeeding, which, because of the survival benefits, remains recommended by World Health Organization (WHO) for at least 12 months even for mothers with HIV.^10,11^ While daily infant antiretroviral prophylaxis is effective at preventing transmission during breastfeeding, long-acting injectable antiretroviral prophylaxis may further reduce vertical HIV transmission because of better adherence.^12,13^ However, given the low risk of vertical HIV transmission with the current standard-of-care, standard randomized trial designs to assess the efficacy of infant long-acting injectable antiretroviral prophylaxis would require sample sizes that may make such trials infeasible to conduct.

As an alternative, we propose an “augmented” clinical trial design that randomizes HIV-exposed infants to a long-acting injectable antiretroviral prophylaxis versus standard-of-care, augmented with observational data under the standard-of-care. This design is inspired by recent work on fusion designs and estimators.^14–16^ Below, we further detail the problems likely to be faced by future trials of long-acting injectable antiretroviral prophylaxis administered to infants. We then present the proposed augmented trial design using a hypothetical example and corresponding simulations. We close with a discussion of advantages and limitations.

## Overview of Vertical HIV Transmission

Oral antiretrovirals can successfully reduce postnatal HIV acquisition,^5,8,10,13,17–21^ with the risk of vertical transmission being as low as 0.9 percent at 24 months when either the mothers receive antiretroviral therapy or infants receive oral antiretroviral prophylaxis.^4,5^ However, real-world challenges remain.^12^ Women with HIV and their infants frequently disengage from care in the postpartum period,^22^ and high maternal viral loads postpartum increase the risk of vertical HIV transmission.^23,24^ Among women initiating efavirenz-based antiretroviral therapy during pregnancy and achieving viral suppression, one-third experienced viral rebound by 1 year postpartum.^12,25^ Preliminary data of women on dolutegravir-based antiretroviral therapy indicate that 21 percent experienced viral loads >1,000 copies/mL at 24 weeks postpartum.^26^

Administering long-acting antiretrovirals to infants as prophylaxis against vertical HIV transmission is a promising alternative to daily oral antiretroviral regimens.^12,13^ In trials of prevention against sexual HIV transmission in adults, long-acting cabotegravir reduced transmission by 66% compared to oral tenofovir disoproxil fumarate/emtricitabine in men and by 88% in women.^27,28^ Recent trial data found no sexual acquisition events among women receiving lenacapavir prophylaxis.^29^ Both agents have existing safety data or ongoing studies in pregnancy and breastfeeding.^30^ For adults, long-acting antiretrovirals offer a reduction in HIV risk while requiring less frequent administration (every 2 months for cabotegravir or every 6 months or longer for lenacapavir).^31^ If the pharmacokinetics allow comparable dosing intervals in infants, then administration of either long-acting antiretroviral could align with existing vaccine schedules during critical periods of recommended breastfeeding.^11,32,33^

## Challenges in Assessing Efficacy of Infant Prophylaxis Against HIV

Given the pathophysiological differences between sexual and vertical transmission of HIV, extrapolation of efficacy from adult populations to pediatric populations may not be acceptable to regulators.^34^ For regulatory approval of a novel agent for use as infant HIV prophylaxis, a registrational trial would require efficacy data in an infant population at risk of HIV acquisition.^34,35^

A key challenge in testing long-acting antiretrovirals for infant HIV prophylaxis lies in a study design and statistical methodology that can detect small differences in infant HIV acquisition, given its low incidence under an effective standard-of-care implemented in a clinical trial setting with sufficient reach and fidelity.^35–39^ In prior trials, maternal antiretroviral therapy or infant antiretroviral prophylaxis throughout breastfeeding resulted in a composite outcome risk of infant HIV acquisition or death as low as 1.8 to 2.6 percent by 12 to 24 months of age.^5,8^ Current standard-of-care is now to provide all mothers antiretroviral therapy for their own health, with daily oral infant postnatal prophylaxis for 6 weeks, then further oral prophylaxis for high-risk breastfeeding infants until maternal viral load suppression or breastfeeding cessation.^1^ Under this standard-of-care, demonstrating that a long-acting antiretroviral agent adds prevention efficacy for infants likely requires outperforming (with sufficient precision) a 12-to-24-month risk of infant HIV acquisition or death of approximately 2 percent or less.

## A Standard Randomized Trial

Suppose we want to conduct a standard randomized trial to assess the prevention efficacy of a candidate long-acting antiretroviral as HIV postnatal prophylaxis administered to infants. Pregnant women would be screened during pregnancy or labor to identify mothers who are (a) presumed or confirmed to be living with HIV and (b) intending to breastfeed. These women would receive the local routine care, including antiretroviral therapy, from enrollment throughout breastfeeding until 24 months postpartum. Mother-infant pairs would be enrolled as soon as possible after birth, with neonates randomized 1:1 to long-acting antiretroviral prophylaxis (throughout breastfeeding, up to 24 months) or standard-of-care prophylaxis (i.e., daily oral antiretroviral[s] for a duration according to local guidelines) and followed for 24 months for the composite outcome of infant HIV acquisition or death from any cause (**Fig 1A**). In this design, assuming 2% risk of the composite outcome in the standard-of-care arm, and 50% risk reduction in the long-acting antiretroviral arm,^8,27,28^ a total sample size of 4,638 mother-infant pairs would be required to achieve 80% statistical power with 5% type 1 error. This sample size, nearly double the total enrolled in the IMPAACT PROMISE trial (*N*=2431),^5^ is likely infeasible to enroll.

**Figure 1.**
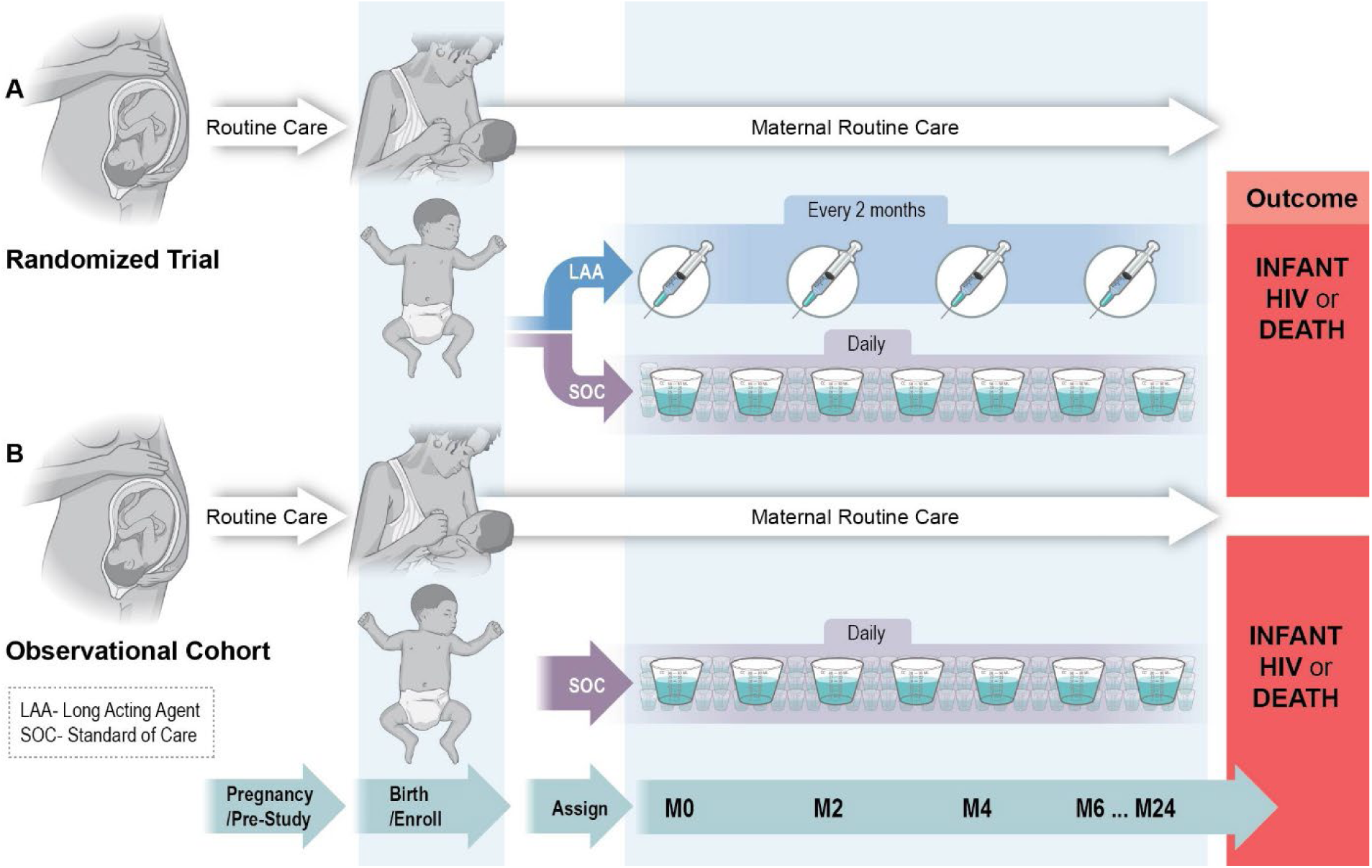
Augmented clinical trial design comparing a novel long-acting injectable agent versus standard-of-care administered to infants. **(A)** In a standard randomized controlled trial (RCT), breastfed infants (born to a woman living with HIV) are allocated at birth to a long-acting agent or standard of care (SOC), then followed for the outcome of infant HIV acquisition or death. **(B)** The RCT’s active control arm is supplemented (or “augmented”) by a concurrent prospective observational cohort, which also follows breastfed infants on SOC from birth for HIV acquisition or death. Combining **(A)** and **(B)** in an augmented clinical trial “fuses” the SOC arms from the RCT and the observational cohort to provide greater precision and study feasibility, as compared to a standard RCT, while preserving validity. Abbreviations: LAA, long-acting agent; RCT, randomized controlled trial; SOC, standard of care.

## Fusion Designs and Augmented Trials

To address the challenge of evaluating infant HIV prophylaxis when transmission incidence is low, we propose an augmented trial, which is premised on a fusion study design combining data from a randomized trial and a concurrent prospective observational cohort (**Fig 1A and B**). Fusion designs integrate data in a principled manner from multiple, possibly heterogeneous datasets, including randomized trials, observational studies, nested two-stage designs, or single-arm studies such as pharmacokinetic or validation studies.^16^ One example of a fusion trial design is a bridged comparison study,^15,40^ which has been previously used to compare adult pre-exposure prophylaxis options across different trials.^41^ The augmented trial design proposed here is novel in that it relies on prospectively collected observational data to supplement the standard-of-care arm. Importantly, relying on a concurrent observational cohort allows investigators to apply the same screening (or eligibility) criteria and variable definitions as the randomized trial, and similarities of settings and geographical footprints help account for temporal and geographic trends.

As with the standard trial described in the previous section, the trial component of this augmented trial would allocate infants with exposure to HIV to either a new long-acting antiretroviral or standard-of-care, albeit with a *k*:1 allocation ratio (where *k*>1). Since regulatory standards would only allow trial participants to receive an investigational agent, the trial component’s asymmetric allocation would allow more balance of participants to the novel agent versus standard-of-care across the combined study. The prospective observational component would simultaneously enroll mother-infant pairs, from similar settings as the trial, who receive the standard-of-care by default and have pre-specified collection of exposure (i.e., maternal and infant antiretrovirals), outcome (infant HIV acquisition or death), and covariates (predictors of cohort selection and outcome).

## Estimators of the Treatment Effect

In both a standard trial and an augmented trial, the intent-to-treat effect can be estimated by comparing the proportion with the composite outcome in the long-acting antiretroviral arm to the same proportion in the standard-of-care arm. However, in the augmented trial, we first convert (i.e., transport) the observational data to mirror the distributions of baseline variables in the trial data, thereby allowing us to validly combine (i.e., pool) the trial and observational standard-of-care arms (**Fig 2**). This “transporting” step is executed by standardizing the observational cohort risk estimates according to the trial’s distribution of prespecified covariates measured at the study baseline (e.g., maternal ART history, maternal viral load, maternal CD4 count).^42,43^ These covariates are chosen based on expert knowledge to include key predictors of vertical HIV transmission,^23,24,44^ whose distributions may differ by the data source (i.e., trial versus observational data). Standardization can be accomplished using a variety of approaches, such as inverse probability weighting, g-computation, or augmented inverse probability weighting (AIPW) — referred to, in this context, as “fusion estimators”.^16,42,45,46^ In the Appendix, we describe an AIPW fusion estimator and provide proofs of consistency and asymptotic normality.

**Figure 2.**
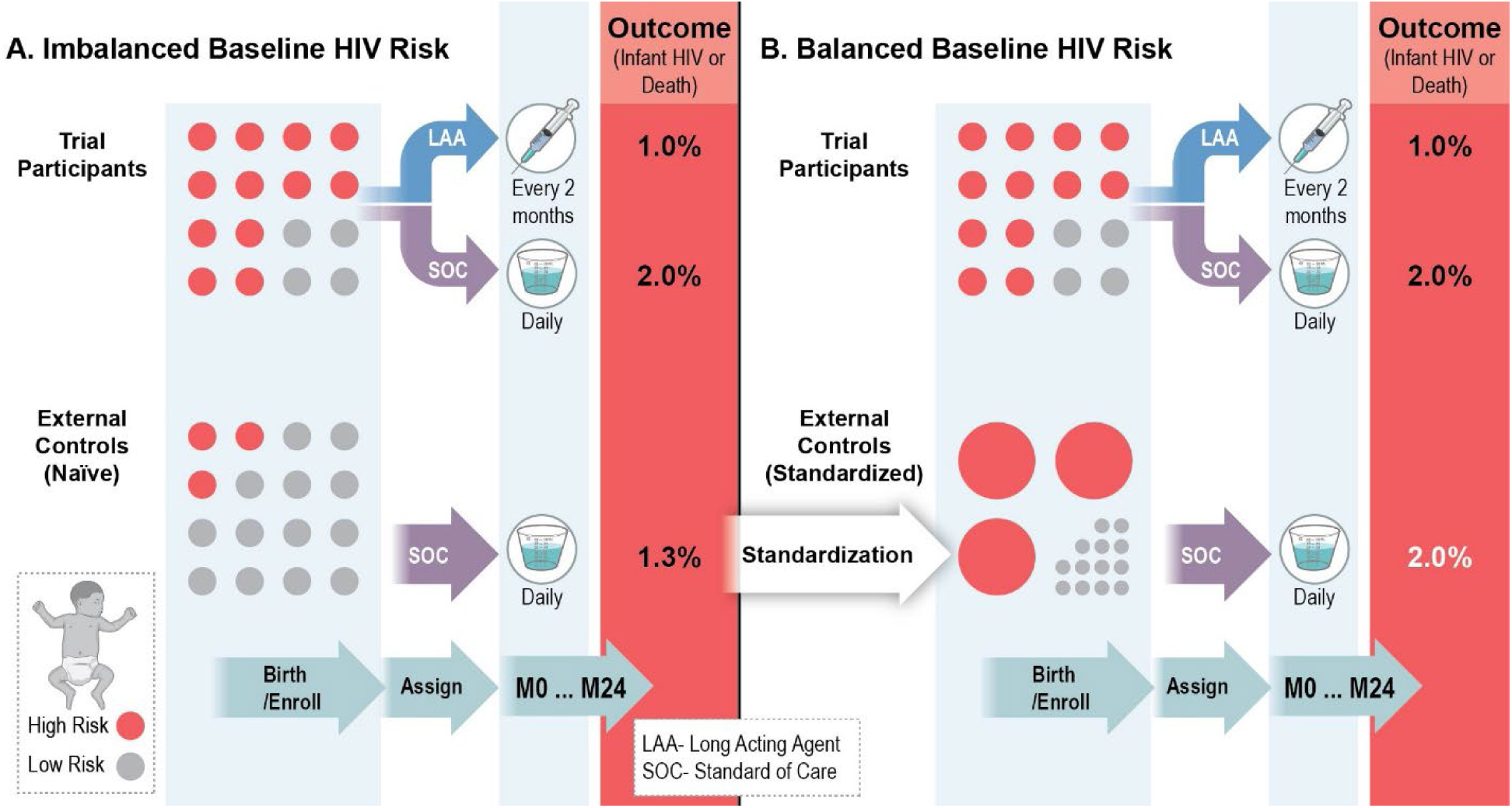
Standardization to account for sampling imbalances in baseline covariates. **(A)** Infants enrolled in the augmented clinical trial may have imbalances in baseline risk of HIV acquisition across the trial participant versus the external control (observational) study populations. In this example, infants at high-risk for HIV acquisition (red dots) are under-represented among the external controls relative to the trial participants (and infants at low-risk [gray dots] are over-represented). These baseline imbalances lead to differences across standard-of-care arms in the observed outcomes of infant HIV acquisition or death. **(B)** With standardization applied to the external controls (note: trial participants remain unchanged), the under-represented infants at high-risk for HIV acquisition are up-weighted (as represented by enlarged red dots), and the over-represented infants at low-risk are down-weighted (as represented by shrunken gray dots). Consequently, the trial participant and external control populations have greater similarity in baseline HIV acquisition risk with corresponding similarity in observed outcomes across the standard-of-care arms. Abbreviations: LAA, long-acting agent; SOC, standard of care.

## A Diagnostic for Pooling

We propose a diagnostic assessment of the validity of pooling (or, “fusing”) trial and observational data, which serves as a check on potential unmeasured sources of bias and missing data assumptions. Specifically, we propose assessing the similarity of the risk of the outcome in the trial and observational standard-of-care arms after the observational data have been standardized to reflect the trial’s distribution of prespecified covariates. This diagnostic is expressed as the difference (or ratio) of risks of the primary composite outcome between the trial standard-of-care arm versus the standardized observational standard-of-care cohort. If these risks are notably different, then the fused estimator is not expected to yield a valid treatment comparison. This approach has previously been applied with external controls^47^ and bridged treatment comparisons,^40,48^ but some challenges remain for practical use of this diagnostic (e.g., the combined sample size must be sufficient for a well-powered test of both the primary composite outcome and the diagnostic, and inference is complicated for pre-test estimators).^49^

## An Illustrative Example

Imagine that the true risk of vertical HIV transmission or infant death were halved by a long-acting antiretroviral compared to the infant standard-of-care prophylaxis.^8,27,28^ Suppose 2,400 mother-infant pairs are enrolled in a trial, randomized 3:1 to infant long-acting antiretroviral or standard-of-care arms, and followed 24 months for the composite endpoint of infant HIV acquisition or death from any cause. An additional 5,000 mother-infant pairs are enrolled into an observational cohort and followed 24 months for the same endpoint. Of the 2,400 trial pairs, 1,600 (67%) are high-risk for vertical HIV transmission. The observational data are composed of fewer high-risk pairs (20%) relative to the randomized data (**Table 1**).

**Table 1:** Illustrative Example of an Augmented Clinical Trial.

| **Data Source** | **Study Arm** | **High Risk**<br>**events / n** | **Low Risk**<br>**events / n** | **Total**<br>**events / n** | **Risk** | **RR**<br>**(95% CI)** | **SE**<br>**(lnRR)** | **SER** |
| --- | --- | --- | --- | --- | --- | --- | --- | --- |
| **Trial Data** | Novel Long-Acting Agent | 12/1200<br>(1%) | 6/600<br>(1%) | 18/1800 | 0.010 | 0.50 (0.24, 1.03) | 0.370 | 1 |
|  | Standard of Care | 10/400<br>(2.5%) | 2/200<br>(1%) | 12/600 | 0.020 |  |  |  |
| **Observational Data** | Standard of Care | 25/1000 (2.5%) | 40/4000<br>(1%) | 65/5000 | 0.013 |  |  |  |
| **Combined Data** | Naïve* | 35/1400<br>(2.5%) | 42/4200<br>(1%) | 77/5600 | 0.014 | 0.73 (0.44, 1.21) | 0.260 | 0.70 |
|  | AIPW* |  |  |  | 0.020 | 0.50 (0.29, 0.86) | 0.276 | 0.75 |

Abbreviations: AIPW, augmented inverse probability weighted estimator; lnRR, natural log of the risk ratio; n, number of participants; RR, risk ratio; SE, standard error; SER, standard error ratio
\* Risks refer to the combined trial and observational standard-of-care arms, while RR and SE refer to comparisons of the novel long-acting agent versus the combined standard-of-care arms.

Suppose a total of 95 endpoints were observed, with 18 endpoints in the long-acting antiretroviral arm (18/1800; 1%), 12 in the standard-of-care arm (12/600; 2%), and 65 in the observational data (65/5000; 1.3%). Here, a naïve combination of the data sources yields a null-biased risk ratio (RR) of 0.73. The AIPW estimator for the combined (i.e., fused) trial and observational data, when the observational data are standardized to the trial distribution of high/low risk, yields an unbiased RR of 0.50. If the trial component were considered a standalone trial, then this “fused” result is similar to the “trial-only” RR of 0.50 but with a 25% smaller standard error (SE). Note also that the 95% confidence intervals (CI) cross the null in the standalone trial and the naïve combination but not in the AIPW-fused study.

Here, the pooling diagnostic compares the risk of infant HIV acquisition or death between standard-of-care groups between the trial and observational cohort. For the naïvely pooled data, this difference was 0.007 (SE = 0.006) with *P* = 0.24. With standardized pooling, the difference was 0.000 (SE = 0.007) with *P* = 1. The diagnostic results suggest that the standardized pooled data provide a more appropriate treatment effect.

## Sample Size and Power for the Augmented Trial

To further explore the properties of the augmented trial design, we conducted a simulation study to explore how statistical power varies by sample size of the trial and observational data, compared to a standard randomized trial. The simulated data were generated following the scenario described above. Power was based on whether the *P*-value for the risk difference (RD) was less than 0.05. Each simulation scenario used 5,000 iterations. Simulations were performed using Python 3.9.6 (Beaverton, OR, USA) (48-51).

For a 1:1-randomized standard trial, power was near 80% for 4,500 mother-infant pairs (**Fig 3A**), aligning with the previous power calculations. Unsurprisingly, an asymmetric allocation of 3:1 had lower power across the evaluated range of sample sizes.

**Figure 3.**
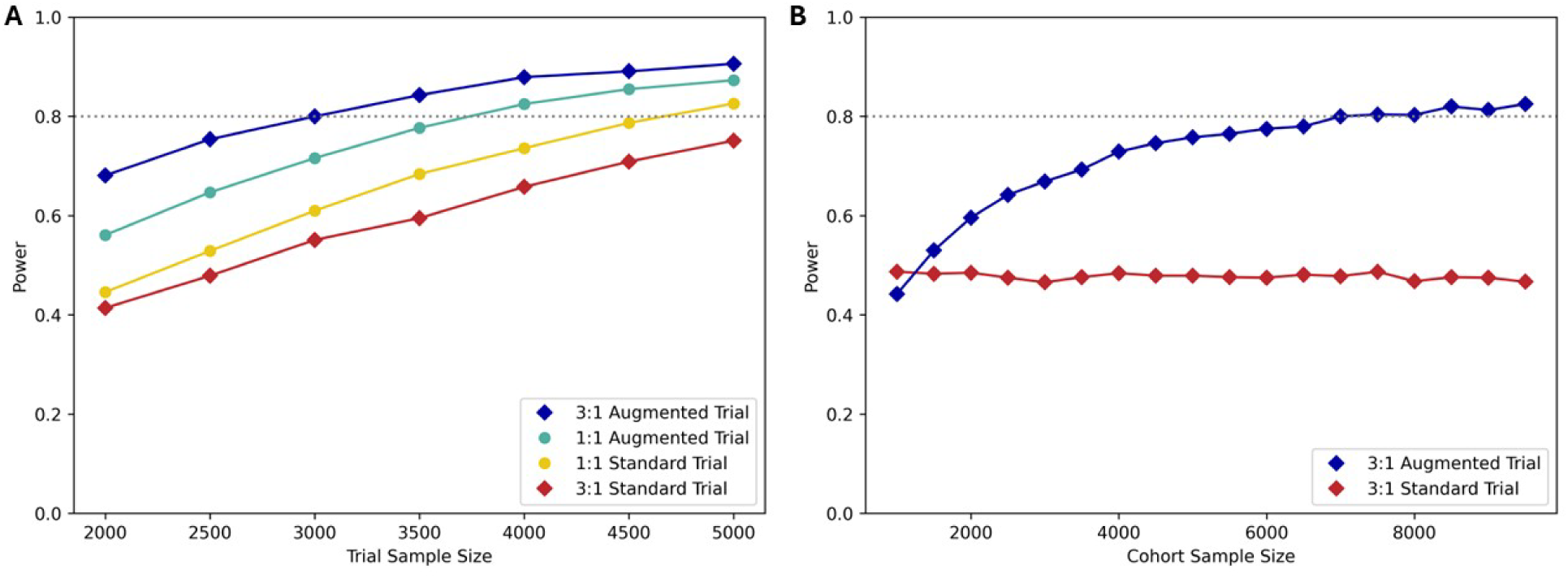
Simulated power of an augmented trial and a standard randomized trial. **(A)** The observational cohort sample size is fixed at 5,000 mother-infant pairs, and power varies according to the trial component sample size by study design and allocation ratio. **(B)** The trial component is fixed at 2,500 mother-infant pairs with 3:1 randomization, and power varies according to the observational cohort sample size by study design. In both panels, the horizontal dotted line denotes the target power of 0.80.

For the augmented trial, we first fixed the observational cohort sample size at 5,000 mother-infant pairs (**Fig 3A**). When the trial component was randomized 1:1, power was above 80% at a trial-component sample size of 4,000 pairs. When the trial component was randomized 3:1, power near 80% could be obtained with as few as 3,000 trial-component pairs, which represents a 33% reduction of the trial component relative to the standard trial. When the trial component was fixed at 2,500 mother-infant pairs (**Fig 3B**), power above 80% was reached with 7,000 or more pairs in the observational cohort component. However, further enrollment did not substantially improve power. As the observational cohort component decreased in size, power decreased, converging on the power of the standard trial as the observational cohort approached 1,500 pairs. This observed lower power of the augmented trial reflects additional uncertainty from standardizing the observational data to the trial’s covariate distribution. Additional performance metrics, including bias, 95% confidence interval coverage, and type 1 error, are reported in the Appendix.

## Financial Costs

With these sample size estimates, what are potential financial consequences? We first assume that the trial component is designed as a phase 2 registrational study, with a primary outcome of safety and a secondary outcome of preliminary efficacy, and conducted according to regulatory requirements such as study product handling, laboratory quality assurance, and site monitoring. We then assume that the observational cohort is of research quality sufficient to meet FDA data standards for regulatory decision-making.^50^ Then, let us assume that central costs are fixed across trial designs and consider per-mother-infant-pair cost scenarios in the trial versus the observational cohort (excluding study product costs) at 5:1 and 10:1, based on conversations with investigators (Personal Communication). To illustrate, we first assign 5 cost units to each trial participant-pair and 1 cost unit to each observational cohort participant-pair. Under the 80% power scenarios above, a standard 1:1-randomized trial with 4,638 mother-infant pairs would total 23,190 cost units. Under the same assumptions for an augmented trial with a 3:1-randomized trial of 2,500 mother-infant pairs and an observational cohort of 7,000 pairs, participant costs would total 19,500 units for a 16% cost reduction. At 10 cost units for trial participant-pairs and 1 cost unit for observational participant-pairs, participant costs would total 46,380 and 32,000 units, respectively, for a 31% cost reduction.

## Discussion

Advances in antiretrovirals for pregnant and lactating women and their infants have dramatically reduced vertical transmission of HIV.^9^ While antiretroviral prophylactic efficacy has benefited infants exposed to HIV, the infrequent transmission poses challenges for future efficacy trials for novel products, especially long-acting antiretrovirals, in an era striving to eliminate pediatric HIV. Under the current standard-of-care, standard randomized trials seem impractical, necessitating innovative designs.^35^ The augmented trial, based on fusion design methodology, offers a potential solution with a sound methodological justification.

### Advantages of the Augmented Trial

In augmented trials, a standard-of-care arm in both the trial and observational data allows for preservation of advantages from randomization while facilitating validation of comparability across trial and observational study populations. We propose using a pre-specified diagnostic to evaluate the comparability of trial and observational data. These design specifications provide an advance over traditional externally controlled studies.

From a regulatory perspective, this design may be particularly advantageous for registrational studies.^51–53^ Regulatory agencies require sufficient safety data to assess benefit versus risk for a study product. The trial component would provide rigorous safety data and reporting, and asymmetric randomization toward the intervention arm should yield greater precision in detecting safety signals, as compared to a 1:1 randomized trial. The United States Food and Drug Administration (FDA) has for decades recognized the value of external controls but has no current guidance regarding trials supplementing a control arm with external controls.^52^ This augmented trial design provides a methodologically sound approach to combining trial and observational data consistent with FDA’s Real-World Evidence Program guidance for study design and data standards.^34,50–54^

Based on our projections, enrolling observational participants under the standard-of-care would provide cost advantages, even when total enrollment in the augmented trial exceeds total enrollment in a standard trial to achieve similar power. Observational studies typically are lower cost endeavors compared to randomized trials, although observational cohorts still require resources for training and data quality assurance. Thus, our projected cost ratios are illustrative based on investigators’ experiences but may vary by site maturity and oversight.

The augmented trial design also improves feasibility. Enrollment of large numbers in trials presents logistical challenges, including finding and screening sufficient eligible participants, supplying and maintaining investigational pharmacies and laboratories, and conducting trial procedures with limited research staff. The estimated sample size requirements under an augmented trial design are within the limits of prior enrollments of comparable studies, e.g. 2,300 to 3,000 total mother-infant pairs in prior clinical trials^5,18,19^ and approximately 2,600 annual (5,000 to 7,000 total over 2 to 3 years) mother-infant patient records in a real-world global observational study.^22^

Further, leveraging external observational data would expedite study enrollment, thereby accelerating study completion, product approval, and ultimately, reaching infants sooner. By example, dolutegravir was initially FDA-approved for adults in 2013 but is still awaiting approval for use in neonates as of October 2025.^55^ Since 120,000 infants acquire HIV annually and experience the highest mortality,^55,56^ each year saved in the research and development pipeline has the potential to prevent many infant HIV acquisitions and thereby impact infant survival.

### Limitations of Proposed Approach and Potential Alternatives

In the proposed augmented trial, sampling (or selection) biases are the principal challenge to external validity and comparability between trial and observational participants.^34,42,57^ Randomization minimizes bias within a trial, but sampling differences across trials and observational cohorts threaten validity. While the proposed AIPW estimator accounts for bias due to measured variables and the pooling diagnostic offers a means to assess residual unmeasured differences across the randomized and observational data, this diagnostic also has drawbacks. If one applies a diagnostic to demonstrate success, then one simultaneously introduces the potential for failure. If the diagnostic suggests important differences are not accounted by standardization, then one might exclude or down-weight the observational data as a way to salvage information.^58^ Alternatively, an interim review could reassess sample size, study duration, and data compatibility, then propose appropriate study modifications. The costs versus benefits of adaptive pooling should be explored further.

Another challenge is data harmonization from multiple data sources. Trials and observational studies are likely to differ by structure, standards, and content.^50,59^ These data sources may also differ in terms of data missingness, measurement differences, and variable categorization. For example, adherence questionnaires capturing any periods of a participant missing all antiretrovirals for at least 7 days could not be reconciled with data on the number of missed antiretroviral doses within 3 days preceding a clinic visit.^60–62^ However, these challenges can largely be avoided by careful protocol planning with a prospective design for the observational component, including planning of data elements (e.g., variables on the intervention, outcomes, and critical covariates for standardization); timing of data collection; and type and categorization of data points. Pilot studies using retrospective data from both data sources would facilitate analysis of key covariates needed, as well as validation of fusion estimators and pooling diagnostics, along with their operating characteristics.

Post-randomization factors, e.g. differential loss to follow-up, may impact data integrity. Although analytical approaches, such as inverse probability of censoring weights,^62–64^ can help to mitigate missing data bias, protocols would benefit from pre-study planning to optimize retention and other mitigation measures across the trial and observational components.

Other analytical approaches and estimators are available, including prognostic scores,^65^ randomization-aware AIPW,^66^ a cross-validated targeted maximum likelihood estimator,^67^ and Bayesian approaches.^58,68–72^ Although we propose one approach, the various options should be examined to determine their relative advantages and disadvantages when applied to a particular clinical scenario.

### Conclusions

Standard randomized trials are likely infeasible for assessing the prophylactic efficacy of newer antiretroviral agents in infants. We propose a novel augmented clinical trial design, where a randomized trial with an active comparator is supplemented with contemporaneous observational data using the same standard-of-care in similar settings. Valid analytical approaches can harmonize sampling differences between study components, and the associated pooling diagnostic can evaluate potential residual biases from unmeasured covariates. Although practical challenges remain, many can be overcome by coordinated study planning and execution. This innovative design provides an opportunity for major advancement toward elimination of infant HIV.

## Appendix

### Setup

Imagine that we have a trial with *n* eligible mother-infant pairs randomized *k*:1 (where *k* ≥ 1) to long-acting antiretroviral or standard-of-care and followed for the composite outcome of infant HIV acquisition or death. To fix ideas, suppose pregnant women aged 18 to 35 years with documented HIV infection are randomized 3:1 to the long-acting antiretrovirals and standard-of-care. The standard arm consists of daily oral infant antiretroviral prophylaxis. In both arms, maternal antiretroviral therapy is given up to 24 months while breastfeeding. In addition, we have observational data on *m* mother-infant pairs under the standard-of-care followed for the same outcome. The observational data are obtained contemporaneously from clinical sites like the sites from which randomized participants were enrolled with the same eligibility criteria as the trial. The interest parameter is the risk ratio, the risk of the outcome under long-acting antiretrovirals divided by the risk under the standard-of-care in the population from which the *n* trial participants were selected.

Let the primary outcome for participant *i* be denoted by *Y_i_*, and long-acting antiretroviral indicated by *A_i_* = 1 (with standard-of-care denoted by *A_i_* = 0). Then the potential outcomes under novel or standard treatment are denoted as *Y_i_*^1^ or *Y_i_*^0^, respectively. Let the randomized trial data be denoted by *S_i_* = 1 (with *S_i_* = 0 for the observational data). Finally, let W*_i_* denote a set of key covariates measured in both data sources (e.g., maternal age, maternal HIV viral load at delivery, maternal CD4 cell count at delivery). We generally suppress the participant index *i* below. The risk ratio, the interest parameter, can be written as *E*(*Y*^1^|*S* = 1)/*E*(*Y*^0^|*S* = 1) = *β*_1_/*β*_0_ = *α*. Next, we show that *α* is identified under a set of assumptions.

Under the above-described study design, *α* is (nonparametrically point) identified given the following three assumptions

- Causal Consistency: *Y_i_^a^* = *Y_i_* if *a* = *A_i_*
- Marginal treatment exchangeability: *Y^a^* ∐ *A* | *S* = 1
- Treatment positivity: Pr(*A* = *a* | *S* = 1) > 0 for *a* ∈ {0,1}

where exchangeability and positivity are given by design. Therefore, it follows that *E*(*Y^a^*|*S* = 1) = *E*(*Y^a^*|*S* = 1, *A* = *a*) = *E*(*Y*|*S* = 1, *A* = *a*) by exchangeability with positivity and causal consistency. Therefore, *α* is identified as it can be expressed using only observable data. A straightforward estimator of this quantity is 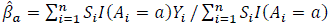, where *I*(*x*) is the indicator function where 1 is returned if *x* is true and 0 otherwise. While identified by design, there can still be substantial imprecision when estimating *α* in practice.

To allow for external data from the observational cohort, the following identification assumptions are added to the previous

- Conditional treatment exchangeability: *Y*^0^ ∐ *A* | W, *S* = 1
- Treatment positivity: Pr(*A* = 0 | W = *w*, *S* = 1) > 0 for *a* ∈ {0,1} and all unique values of *w* in the *S* = 1 population
- Conditional sampling exchangeability: *Y*^0^ ∐ *S* | W
- Sampling positivity: Pr(*S* = 0 | W = *w*) > 0 for *w* where Pr(*S* = 1 | W = *w*) > 0

The first two of these assumptions are also given by design in marginally randomized trials when W occurs prior to randomization. The latter two assumptions are commonly used in the transportability literature.^42,73^ Given these assumptions, it follows that

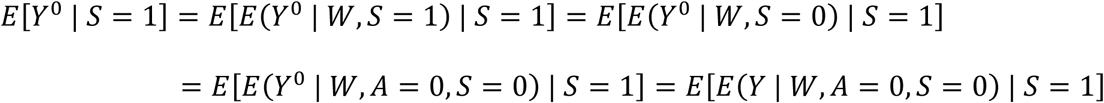

via the law of iterated expectations, sampling exchangeability with positivity, that *A* = 0 when *S* = 0, and causal consistency. This result shows how the observational cohort data can be used to supplement the standard-of-care arm under the previous identification assumptions.

### Fusion Estimator

We present augmented inverse probability weighted (AIPW) estimator of *α* that leverages the observational data. In the AIPW estimator considered below, *β*_1_ is estimated using only the trial data from the novel arm with 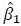. The AIPW estimator then attempts to improve precision by estimating *β*_0_ using the standard-of-care arm of the trial augmented with the observational data. Importantly, the observational data is standardized using a sampling model and outcome model.

To describe the AIPW estimator and prove its asymptotic properties, we rely on estimating equation theory. An estimating equation is defined as 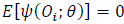, where *ψ* is a finite-dimension estimating (vector) function, *O_i_* is the observed data, and *θ* is a finite-dimension parameter vector.^74^ The corresponding estimators, 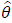, have also been referred to as M-estimators or Z-estimators. Here, the AIPW estimator is based on a weighted regression implementation, which is expected to be more efficient than the standard canonical AIPW estimator.^75,76^ The stacked estimating functions are

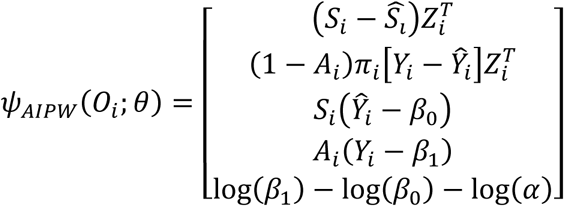

where 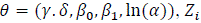 is the design matrix (here, 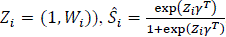, 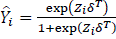, and 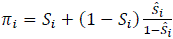. Here, the first estimating function is a logistic regression model for the sampling probability and the second is a weighted logistic regression model for the outcome restricted to only those with *A_i_* = 0 (this means *A_i_* does not need to be included in *Z*). Note that while the design matrix is the same for both the outcome model and sampling model, this need not be the case. The third estimating function is the predicted risk under the standard of care from the outcome model predictions among the trial observations. The fourth estimating function is the predicted risk under the novel treatment and is based on the usual estimator for *β*_1_.The final estimating function is for the (log-transformed) risk ratio. Note that the risk difference can be estimated by using the identity function instead of the log function for the *β*’s in the final estimating function, which would become *β*_1_ − *β*_0_ − *α*.

### Proofs for the Fusion Estimator

We show that the AIPW estimator is consistent and asymptotically normal by demonstrating each of the estimating equations in unbiased. Note that the sampling model in the first estimating function is unbiased following standard maximum likelihood theory. Now consider the second estimating function in *ψ_AIP_*_W_. First, note that

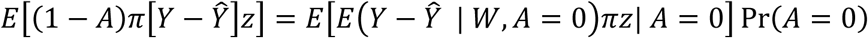

for all *z* ∈ *Z*. Focusing on the inner expectation, it follows from correct specification of the outcome model that

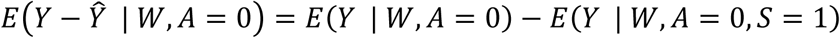

This difference is equal to zero following the identification assumptions for transporting. Therefore, this estimating equation is unbiased by the zero property of multiplication. Now consider when the sampling model is correctly specified. Here, *z* = 1 to simplify exposition without a loss of generality. Further, we drop (1 − *A*) since this term is only meant to make the correct specification of the outcome model more plausible in practice (i.e., one could instead include *A* in *Z* to borrow more information when fitting the outcome model). Therefore,

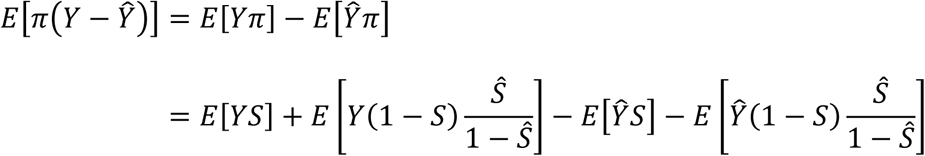

following from the definition of *π*. Consider the second summand, it follows that

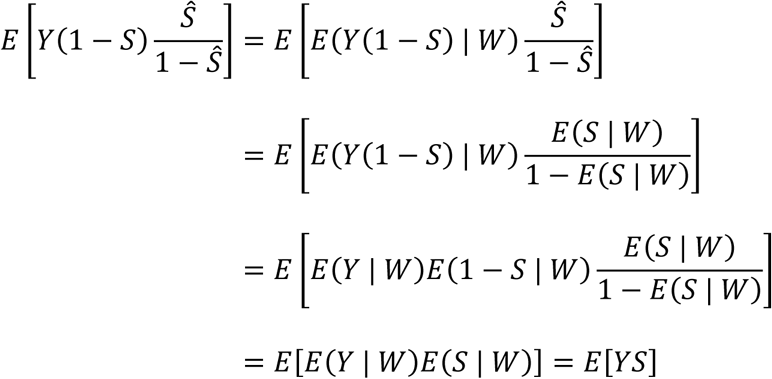

via iterated expectation by W and independence, correct sampling model specification, sampling exchangeability, and algebraic manipulation. The last summand is equal to

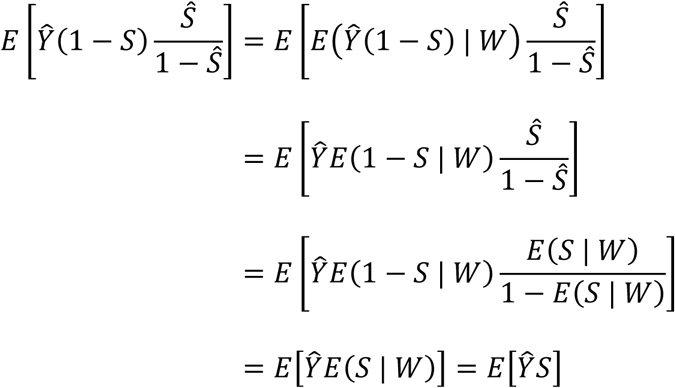

via iterated expectation by W and independence, correct model specification, and cancellation of terms. Putting these summands together, it follows that

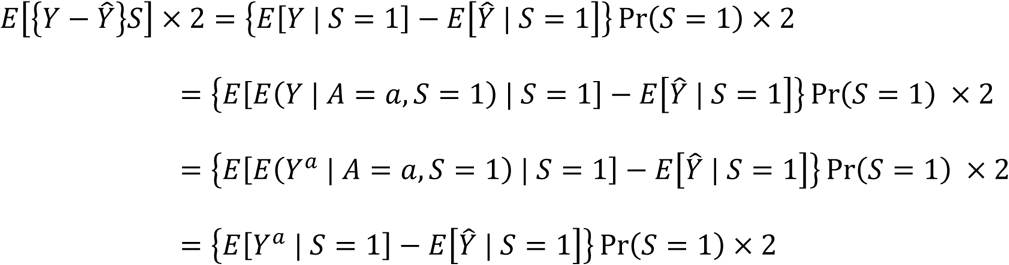

following from manipulations of the expectations, iterative expectation by *A*, causal consistency, and undoing the iterative expectation by *A*. Therefore, the solution to the estimating equation requires that the outcome model is equal to *E*[*Y^a^* | *S* = 1] regardless of whether it is correctly specified (besides inclusion of an intercept and main term for *A*). Therefore, the second estimating equation is unbiased as long as one of the two nuisance models is correctly specified. Given this result, it directly follows that the third estimating function is unbiased as long as one of the two models is correctly specified. For the fourth, it follows from algebraic manipulation that *E*[*A*(*Y* − *β*_1_)] in unbiased from the estimator presented with the identification results.

Finally, the fifth estimating function is unbiased directly following the definition of each of the parameters.

Given suitable regularity conditions for non-identically distributed data,^77^ it follows that as 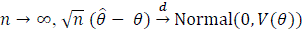, where the variance is 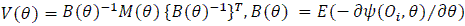, and 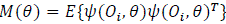. The variance can then be estimated using the empirical sandwich variance estimator. Here, the fusion AIPW estimator is doubly robust for point estimation and the empirical sandwich variance estimator is doubly robust for variance estimation with respect to the sampling and outcome models.^78^

### Simulations

Next, we explore some finite sample properties of these AIPW estimators in a proof-of-concept simulation experiment. We present two scenarios of simulations. In both scenarios, there are 2,400 trial participants randomized 3:1 to the novel or standard arms, and 5,000 observational participants all under standard treatment. Also in both scenarios, 2/3 of trial participants and 1/5 of observational study participants are high-risk (i.e., W = 1).

The first scenario was the same as described in the paper, with the risk of outcome in the trial is 1% except for high-risk participants in the standard arm, where it is 2.5%. The risk of outcome in the observational data is 2.5% for high-risk participants and 1% for low risk. In the second scenario, the risk of outcome in the trial is 5% except for high-risk participants in the standard arm, where it is 12.5%. The risk of outcome in the observational data is 12.5% for high-risk participants and 5% for low risk. In both scenarios the true parameter of interest is a risk ratio of 1/2.

In addition to the AIPW estimator, the estimator that uses only the trial data and a naïve fusion estimator that does not standardize the observational data are considered. For 5,000 different data sets, we report bias (defined as the mean of the estimated minus the truth), relative bias (defined as bias divided by the true parameter), the empirical standard error (ESE; defined as the standard deviation of the estimates), the standard error ratio (SER; defined as the average of the estimated standard errors divided by the ESE), 95% confidence interval (CI) coverage (defined as the proportion of confidence intervals containing the true value), and rejection (defined as the proportion of 95% CIs that did not include the null).^79^ Results are reported for both the risk difference and the (log-transformed) risk ratio. Simulations were conducted in Python 3.9.6 (Beaverton, OR, USA) with the following packages: NumPy,^80^ SciPy,^81^ pandas,^82^ delicatessen,^83^ and matplotlib.^84^

**Table S1** presents the simulation results for the first scenario. For the risk difference, bias was near zero for all estimators. However, this small amount of bias for all estimators is likely due to the event being rare, as indicated by the relative bias results. A discrepancy between estimators is most apparent in the 95% CI coverage, where the naïve fusion estimator had below 40% coverage. Both the trial-only and fusion AIPW estimators had coverage near the expected level of 95%, but the fusion AIPW had a smaller ESE. The precision benefit of the observational cohort is most apparent in the rejection, where the fusion AIPW had near 80% power, more than double the trial-only estimator. Patterns were similar for the risk ratio, but bias was more pronounced for the naïve fusion. Further, there appeared to be some finite-sample bias for both the trial-only and AIPW estimators of the risk ratio.

**Table S1.** Simulation results for the first scenario.

|  | Bias | Relative bias | ESE | SER | Coverage | Rejection |
| --- | --- | --- | --- | --- | --- | --- |
| Risk Difference |  |  |  |  |  |  |
| Trial-Only | 0.000 | 0.005 | 0.006 | 0.98 | 93.3% | 33.2% |
| Naïve Fusion | 0.006 | -0.622 | 0.003 | 1.00 | 38.2% | 29.4% |
| Fusion AIPW | 0.000 | 0.009 | 0.004 | 1.01 | 95.1% | 77.1% |
| Risk Ratio |  |  |  |  |  |  |
| Trial-Only | 0.014 | 0.029 | 0.402 | 0.96 | 95.5% | 46.2% |
| Naïve Fusion | 0.350 | 0.700 | 0.273 | 0.98 | 68.8% | 21.3% |
| Fusion AIPW | 0.009 | -0.047 | 0.287 | 0.99 | 95.7% | 74.2% |

AIPW: augmented inverse probability weighting, ESE: empirical standard error, SER: standard error ratio

For the second scenario, the finite-sample bias present in the previous scenario disappeared (**Table S2**). Both the trial-only and fusion AIPW estimators had near zero bias and near nominal 95% CI coverage for both parameters. As expected, the naïve estimator was biased.

**Table S2.** Simulation results for the second scenario.

|  | Bias | Relative bias | ESE | SER | Coverage | Rejection |
| --- | --- | --- | --- | --- | --- | --- |
| Risk Difference |  |  |  |  |  |  |
| Trial-Only | 0.000 | -0.001 | 0.013 | 0.98 | 94.2% | 97.1% |
| Naïve Fusion | 0.031 | -0.625 | 0.006 | 1.02 | 0.0% | 85.6% |
| Fusion AIPW | 0.000 | 0.000 | 0.008 | 1.01 | 95.1% | 100% |
| Risk Ratio |  |  |  |  |  |  |
| Trial-Only | 0.003 | 0.006 | 0.162 | 0.99 | 94.7% | 98.1% |
| Naïve Fusion | 0.370 | 0.740 | 0.113 | 1.02 | 11.1% | 82.9% |
| Fusion AIPW | -0.004 | -0.007 | 0.119 | 1.02 | 95.3% | 100% |

AIPW: augmented inverse probability weighting, ESE: empirical standard error, SER: standard error ratio

### Type 1 Error

To explore Type 1 error, the first simulation scenario was repeated except that the risk of outcome in the trial is 1% for low-risk participants and 2.5% for high-risk participants, regardless of the assigned treatment. Here, the observational cohort size was fixed at 5,000 while the size of the trial was allowed to vary from 2,000 to 5,000 in increments of 500. Further, results were compared for 1:1 and 3:1 randomization using the trial-only (i.e., standard trial) and fusion (i.e., augmented trial) AIPW estimators.

As seen in **Figure S1**, the proportion of 95% CIs that excluded the null was near 0.05 for all trial designs across the sample sizes considered. These results indicate that the Type 1 error rate was maintained for all study designs with a rare outcome.

**Figure S1.**
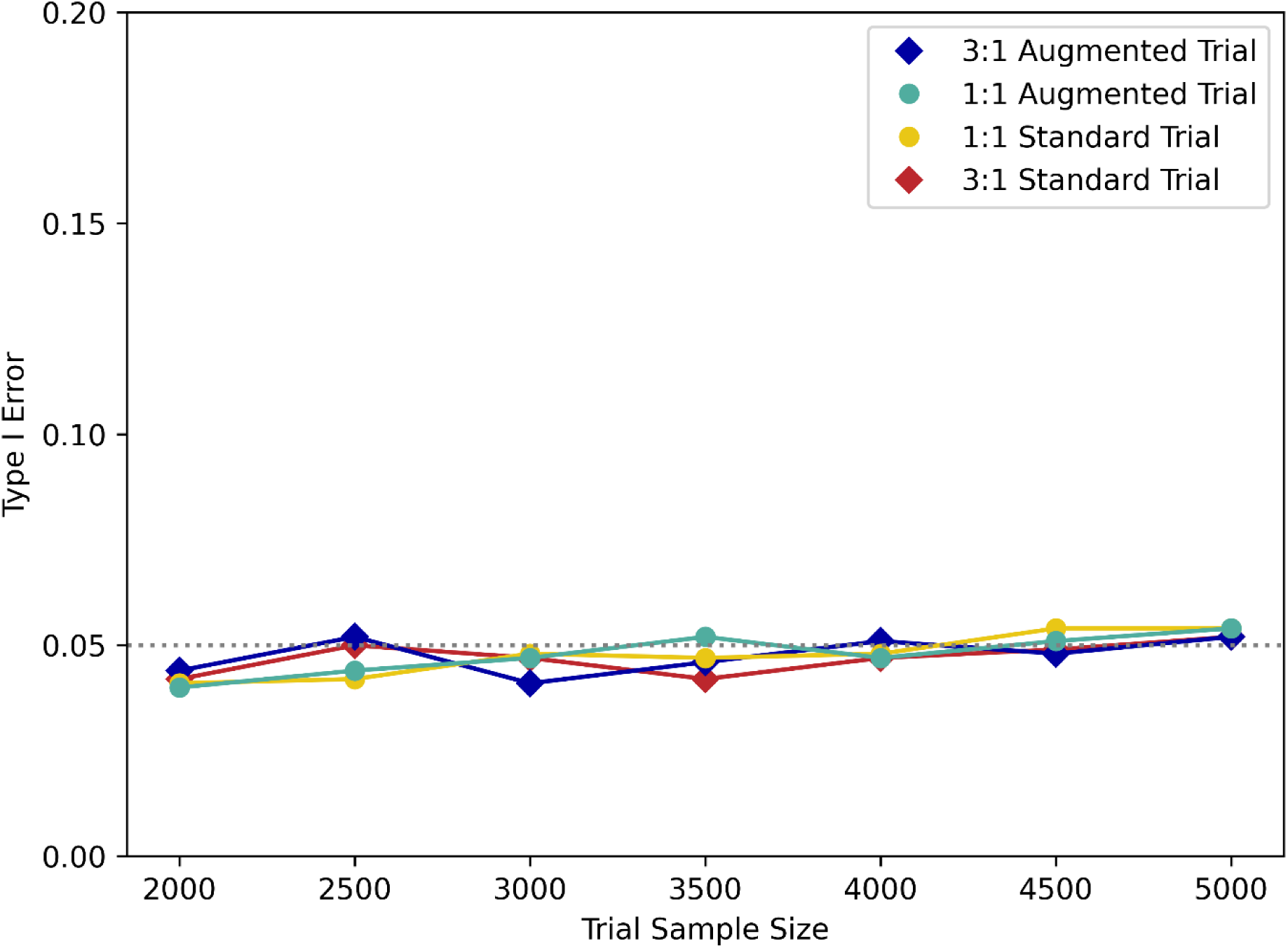
Type 1 error by trial sample size. The observational cohort component sample size is held constant at 5,000 mother-infant pairs. Type 1 error is plotted according to the trial component sample size by study design and allocation ratio. The horizontal dotted line denotes a type 1 error of 0.05.

## Acknowledgments

We would like to acknowledge Ryan Kissinger, Anita Mora and Austin Athman from the Visual and Medical Arts unit at NIAID for their expertise in creating the illustrations. We thank Issa Dahabreh, M.D., Sc.D., M.S., for reviewing and providing comments on this manuscript.

## Disclaimer

The opinions expressed in this article are the authors’ own and do not necessarily reflect the views of the National Institutes of Health, the Department of Health and Human Services, or the United States government.

## Data Availability

Code is available at https://github.com/pzivich/publications-code.

